# Prediction of Unplanned Hospital Readmission using Clinical and Longitudinal Wearable Sensor Features

**DOI:** 10.1101/2023.04.10.23288371

**Authors:** Haben H. Yhdego, Arshia Nayebnazar, Fatemeh Amrollahi, Aaron Boussina, Supreeth Shashikumar, Gabriel Wardi, Shamim Nemati

## Abstract

Predictive models have been suggested as potential tools for identifying highest risk patients for hospital readmissions, in order to improve care coordination and ultimately long-term patient outcomes. However, the accuracy of current predictive models for readmission prediction is still moderate and further data enrichment is needed to identify at risk patients. This paper describes models to predict 90-day readmission, focusing on testing the predictive performance of wearable sensor features generated using multiscale entropy techniques and clinical features. Our study explores ways to incorporate pre-discharge and post-discharge wearable sensor features to make robust patient predictions. Data were used from participants enrolled in the AllofUs Research program. We extracted the inpatient cohort of patients and integrated clinical data from the electronic health records (EHR) and Fitbit sensor measurements. Entropy features were calculated from the longitudinal wearable sensor data, such as heart rate and mobility-related measurements, in order to characterize time series variability and complexity. Our best performing model acheived an AUC of 83%, and at 80% sensitivity acheived 75% specificity and 57% positive predictive value. Our results indicate that it would be possible to improve the ability to predict unplanned hospital readmissions by considering pre-discharge and post-discharge wearable features.

## Introduction

Hospital readmissions are becoming increasingly more relevant due to the significant financial burden for public and private payers [1]. In the United States, annual costs for hospital readmission reach $41.3 billion, making it one of the costliest events to treat nationwide [2]. Studies estimate that some of these hospital readmissions are avoidable [3], so identifying potential hospital readmissions early and providing better post-discharge planning is crucial [4]. As a result, there is a growing interest by healthcare providers and insurers to reduce readmission risks by determining the probability of hospital readmissions in the future.

Machine Learning (ML) has demonstrated useful potential with the rise of clinical data in predictive modeling for healthcare by using the capability of discerning unique patterns in data without being explicitly programmed. Machine learning models have been implemented to improve clinical decision-making through different data modalities, such as structured and unstructured clinical data in electronic health records (EHRs) and medical imaging [5]. The ability of ML models to handle multivariate and multimodal data allows for their potential application in forecasting hospital readmissions using a broader set of data modalities and features.

Michailidis et al. used administrative/demographic variables, medical-clinical variables, and operational status data from the General Hospital of Komotini, Greece, to predict hospital readmissions [6]. With a Random Forest classifier, their model achieved an area under the curve (AUC) of 0.78. Mahmoudi et al. evaluated 41 studies on predictive modeling compared to statistical methods for hospital readmissions and found that machine learning techniques generally performed better [7].

The use of wearable technology has recently grown in popularity for health monitoring. Wearables use different sensors, such as temperature, accelerometers, optical, and biometric sensors, to track various physiological attributes [8]. Other attributes like monitoring activity levels may also indicate future clinical outcomes. As a result, wearable technologies are increasing applied to building of healthcare solutions. Some of these include clinical decision-making, prevention of diseases, and maintenance of health [9].

In this work, we make use of multimodal and multidimensional data from a publicly available diverse multi-institutional dataset to predict unplanned hospital readmission. We integrate EHR data with wearable data to develop a machine learning model to predict 90-day hospital readmissions and assess the added impact of wearable features. This study demonstrates the potential for wearable technology to enhance the data used in predictive models for hospital readmissions. It may also suggest wearable data’s promise in future studies for predictive modeling in other clinical domains requiring post-acute and long-term monitoring.

## Methods

### Study Cohort

The All of Us program is a longitudinal cohort study by the National Institutes of Health (NIH) aimed to advance precision medicine and improve health by partnering with up to one million diverse participants nationwide [10, 11, 12]. The creation of the All of Us multi-center cohort provides a research database that combines participant-derived survey information, physical measurements, EHRs, biospecimens, and wearables [13]. All of Us Researcher Workbench, a cloud-based analytical platform built by the program for approved researchers, was used to access the dataset. We used a retrospective longitudinal cohort of participants from the All of Us Controlled Registered Tiered Dataset v6 CDR (C2022Q2R6) [11, 14, 12, 15].

### Patient Selection from All of Us Database

Patients are selected from the All of Us database that satisfy the following criteria:

1. Patients admitted to hospitals as Inpatient via emergency departments (ED), general wards, or intensive care units (ICUs) and stayed in the hospital for at least 48 hours.
2. Patients that wore Fitbit sensors and were at least 18 years old.
3. Availability of minute-level heart rates and movement-related data, to enable derivation of multiscale entropy features for logitudinal predictions.

According to criteria 1-2, admissions for short-duration procedures and so forth without Fitbit wearable sensors were removed, resulting in 12,844 patients. After that, criteria 3 was applied to find patients with minute-level data, yielding 11,575 patients. Lastly, the above three criteria were applied to define initial admission as the first hospitalization for each patient. Nine hundred seventy-six patients were identified that fulfilled those criteria. Of 976 patients, 25 and 37 were readmitted to any partner hospitals in the All of Us program within 30 days and 90 days of the index admission, respectively. The characteristics of the Fitbit Inpatients participants are found in Table 2.

**Table 1:** Characteristics of the Fitbit Inpatient Participants.

| Variables | Not readmitted within<br>90 days(n=939) | Readmitted within<br>90 Days(n=37) |
| --- | --- | --- |
| Age<br>[ IQR ] | 52<br>[ 37 – 64 ] | 56<br>[ 49 – 65 ] |
| Gender (Female %) | 70 | 67 |
| Median Number of Steps<br>[ IQR ] | 5720<br>[3345 – 8436 ] | 2481<br>[ 1018 – 4636 ] |
| Median Activity Calories<br>[ IQR ] | 878<br>[ 589 – 996 ] | 545<br>[ 291 – 889 ] |
| Median Length of Hospital Stay (LOS)<br>[ IQR ] | 17<br>[ 3 – 48 ] | 72<br>[ 28 – 144 ] |

**Table 2:** List of features and their descriptions.

| Name | Unit of Measurement | Name | Unit of Measurement |
| --- | --- | --- | --- |
| **Demographics** |  |  |  |
| Age | - | Hospital Stay | hour |
| **Wearable Features (Measured daily)** |  |  |  |
| Heart rate:<br>Minimum<br>Maximum | beats/minutes | Activity Calories | - |
| Calories BMR | - | Calories out | - |
|  |  | Marginal calories | - |
| Activity level:<br>Sedentary<br>Fairly_active<br>Lightly_active<br>very_active | Minutes | Sum steps | - |
| **Derived Wearable Features**<br>**(Measured minute-level)** |  |  |  |
| Heart rate:<br>Multiscale entropy (MSE)<br>Average Heart Rate | Intraday Steps:<br>Average number steps | - | - |
| **Clinical Features** |  |  |  |
| Base Excess | mmol/L | Serum Glucose | mg/dL |
| Fraction of inspired Oxygen | % | Lactate | md/dL |
| pH | - | Magnesium | mmol/dL |
| Partial Pressure of CO2 | mmHg | Phosphate | mg/dL |
| Oxygen Saturation | % | Potassium | mmol/L |
| Aspartate transaminase | IU/L | Total Bilirubin | mg/dL |
| Blood Urea Nitrogen | mg/dL | Troponin I | ng/mL |
| Alkaline Phosphate | IU/L | Hematocrit | % |
| Calcium | mg/dL | Hemoglobin | g/dL |
| Chloride | mmol/L | Partial thromboplastin time | Seconds |
| Creatinine | mg/dL | White blood count | Count *10<sup>3</sup> /μL |
| Bilirubin direct | mg/dL | Fibrinogen | mg/dL |
| c-reactive protein | mg/L | Platelets | Count *10<sup>3</sup> /μL |
| **Vital Signs (Dynamical Features)** |  |  |  |
| Heart rate | beats/minutes | Diastolic BP | mmHg |
| Temperature | degC | Respiration rate | Breaths per minutes |
| Systolic BP | mmHg | - | - |

The 90-day period for unplanned readmission was chosen to include a broader class of at risk patients, and to further power the study with a total of 37 readmitted patients with wearable data. Readmissions were only counted for hospital stays that were at least 48 hours long.

### Features

Fitbit’s data of AllofUs is distributed across four tables representing two main data elements: heart rate and activity data summarized as a daily reports. Additionally, heart rate minute level and intraday steps contain heart rate and activity data represented granularity at the minute level. For every patient, we queried the sum of daily steps, minimum heart rate, maximum heart rate, activity calories, Basal Metabolic Rate (BMR) calories, calories out, marginal calories, elevation, sedentary minutes, “fairly” active minutes, “lightly” active minutes, and “very active” minutes from heart rate and activity daily summary tables.

Besides using the different features from the daily report, we also calculated features like multiscale entropy, average heart rate, and the average number of steps from the minute-level data. Multiscale entropy methodology has been applied to analyze the complex behavior of heart rate variability [16]. Multiscale entropy analysis calculates an entropy rate over different time scales to assess the complexity of a time series. Figure 1 shows the average heart rate of readmitted and non-readmitted patients over the cohort study period. Figure 2 shows the daily entropy of the minute-level heart rate data for a cohort of hospitalized patients, including pre-discharge (day 1 through day 9) and post-discharge (day 9 through day 15), separately for readmitted and non-readmitted patients.

**Figure 1:**
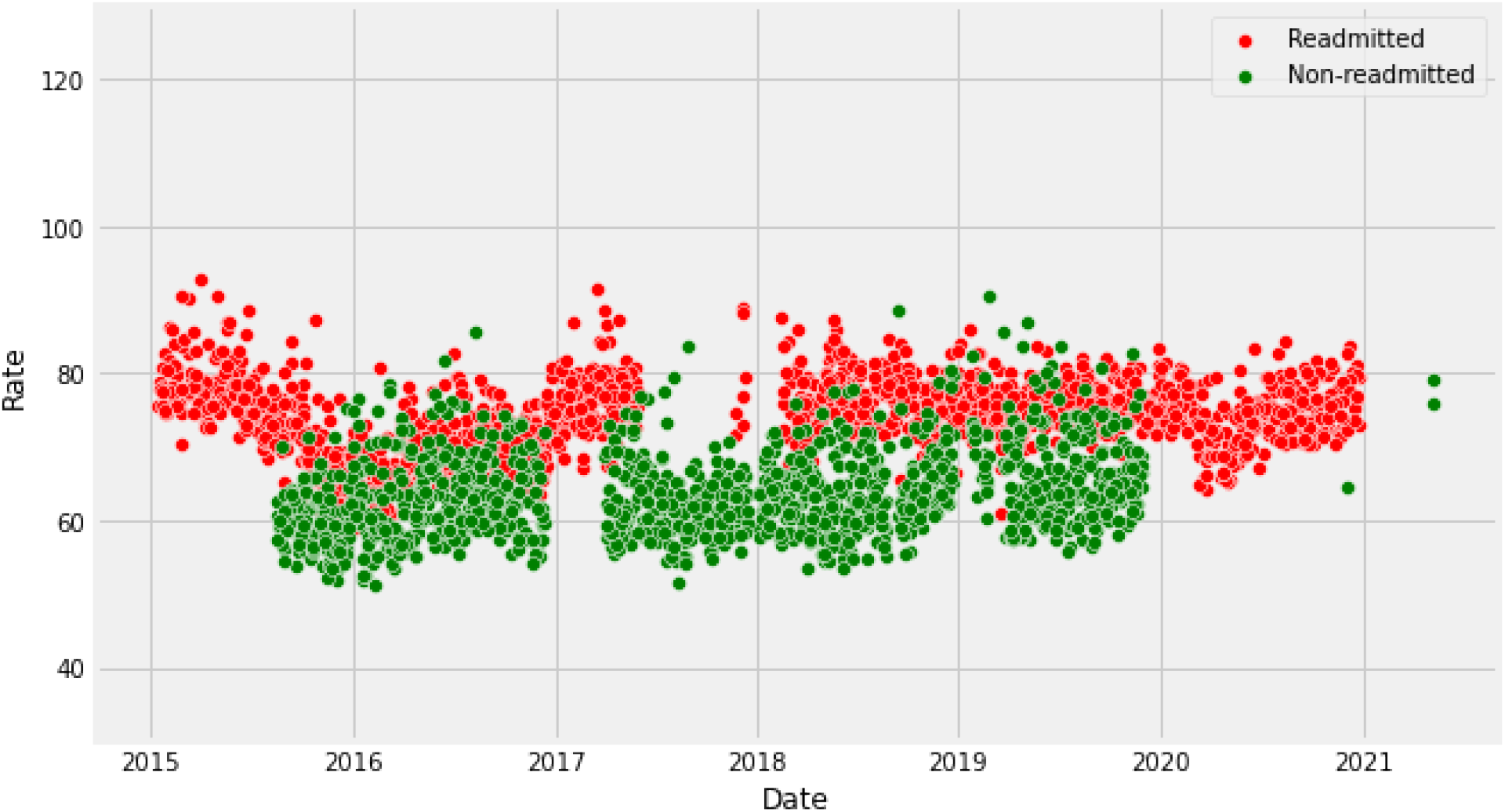
Sample average heart rate feature derived from the minute level data for admitted and non-readmitted.

**Figure 2:**
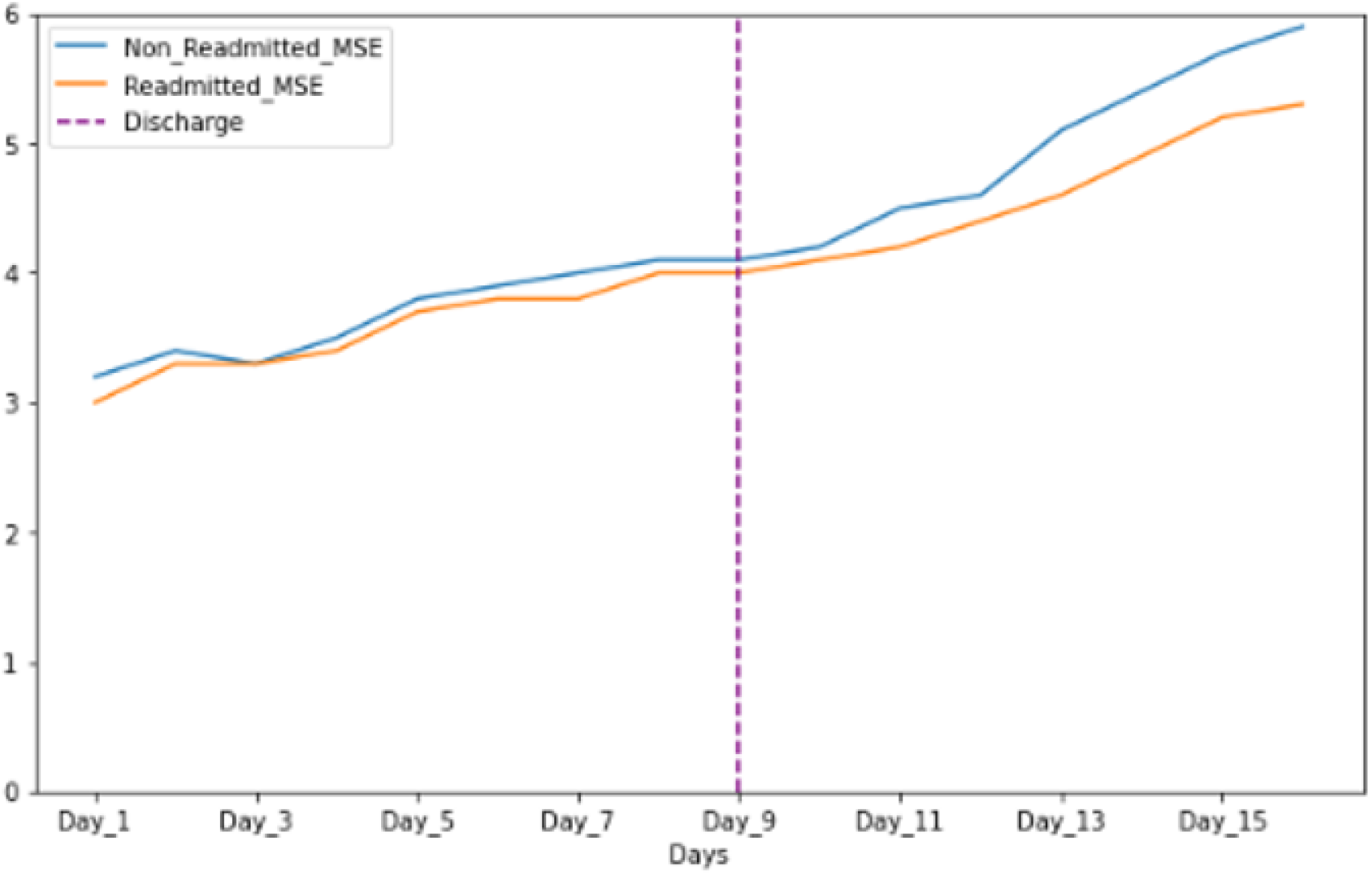
Multiscale entropy for re-admitted and non-readmitted patients at Scale 7 that is calculated from the heart rate minute-level data.

Our model uses around 100 features, including length of stay and age, around 60 clinical features (laboratory results, vitals, and demographic features), and 40 wearable features. Clinical characteristics were queried from the AllofUs dataset and were selected to represent the most commonly measured lab data. Wearable data was extracted from patients who shared their Fitbit data with All of Us. A list of our features can be found in Table 1.

### Statistical Methods

The 5th, 50th, and 95th percentiles for each wearable feature were taken during the index admission, prior to discharge, after discharge until the readmission date or up to 90 days. Furthermore, for the laboratory measurements that were extracted using concepts represented in Observational Medical Outcomes Partnership (OMOP) common data model (CDM), we used each laboratory measurement’s 5th, 50th, and 95th quantiles. Missing values for the continuous features were imputed with the mean, and KNN imputation was used for categorical and binary features. At a fixed 80% sensitivity level, all prediction results are based on a 5-fold cross-validation system (80% training and 20% testing). The area under receiver operating characteristic (AUROC) curves is reported, as well as specificity and positive predictive value (PPV). We combined all the testing set predictions across all the five folds to report a single pooled AUROC [17].

### Algorithm Model

We trained a gradient-boosting model to predict unplanned 90-day readmission with a 48-hour rolling window for inpatients with clinical and wearable features. To predict hospital readmission, we considered four separate models based on the following: 1) Pre-discharge clinical and demographics data, 2-3) Pre-discharge and post-discharge daily summary Fitbit data, and 4) combined features from models 1-3 with entropy features derived from the minute-level Fitbit data. All four models were classified using a gradient-boosting classifier, where internal cross-validation (i.e., within the training set) was conducted to determine the optimal regularization.

## Results

The resulting cohort included 976 inpatient subjects with wearable data, where 37 patients were readmitted to the hospital. Using stratified 5-fold cross-validation, our 90-day model that includes clinical and wearable features achieves an AUC of 0.83. Without the clinical variables and only using the pre-discharge and post-discharge wearable features, the 90-day model achieves an AUC of 0.74. We also ran a separate model by augmenting the wearable data with derived features during the hospital, after discharge, and for both the hospital duration and after discharge. Including derived features improves the wearable sensor model performance to a median AUC of 0.76. We performed feature selection for the final model to reduce overfitting. These results can be found in Table 3.

**Table 3:** Performance Metrics.

| Models | AUC | Specificity | PPV |
| --- | --- | --- | --- |
| Pre-charge clinical features and Demographic patient features | 0.73 | 0.56 | 0.36 |
| Pre-charge and Post-discharge daily summary Fitbit data | 0.74 | 0.61 | 0.42 |
| Combining features from model 2 with Multiscale Entropy features derived from the minute-level Fitbit data | 0.76 | 0.71 | 0.52 |
| Combining features from model 3, and model 1 | 0.83 | 0.75 | 0.57 |

## Discussion

Our results demonstrate that features extracted from wearable sensors is a promising category of health data in the development of predictive models for hospital readmission. Wearable features improve the prediction performance of a model that predicts 90-day readmission with clinical variables. To the best of our knowledge, this work is among the first attempts to combine wearable and clinical features to predict unplanned hospital readmission. The addition of wearable features increased the AUC and positive predictive value of the model in comparison to the model that used only clinical variables. This indicates that the wearable data, from fitness monitors and activity trackers such as smartwatches, includes helpful information for predicting readmission. Therefore, future readmission models may consider using longitudinal wearable data such as minute-level heart rate and accelerometry to improve their predictive capabilities. Moreover, our analysis showed that entropy was higher after patient discharge, and entropy in non-readmitted patients was slightly higher than in admitted patients. Hence, analysis of longitudinal data using techniques such as multiscale entropy analysis may help to characterize readmitted patients and distinguish them from non-readmitted patients.

The All of Us dataset used in this study is among the most diverse data sources available for retrospectively analysis, and presents a unique opportunity for building generalizable predicitive models. Some limitations of the study include the relatively small cohort size (976) and the imbalanced number of readmitted patients (37). A more significant number of All of Us participants with wearable data in the future will help improve the model’s performance. Our future work involves locally validating this model at UC San Diego in a prospective observational study.

## Conclusion

With the help of wearable sensor features, it is possible to predict hospital readmission within 90 days. The prediction result was improved by incorporating multiscale entropy-based measures of HR dynamics and other data derived from wearable sensors. Additionally, adding clinical features to the model greatly improved model performance. Prospetive studies in collaboration with population health management teams including case-wrokers are needed to devise actionable interventions for reducing unplanned hospital readmissions.

## Data Availability

The AllofUs cohort is publicly available at www.allofus.nih.gov.

## Acknowledgments

S.N. is funded by the National Institutes of Health (R35GM143121). He is co-founder of a UCSD start-up, Healcisio Inc., which is focused on commercialization of advanced analytical decision support tools. G. W. is funded through an early career award from the National Institute of General Medical Sciences (K23GM146092).

## Notes

### Competing Interest Statement

The authors have declared no competing interest.

### Author Declarations

This was a retrospective multicenter cohort study consisting of participants in the AllofUs (Registered Tired Dataset v4 CDR data dictionary [R2020Q4R3]) longitudinal study. Institutional Reviewing Board (IRB) approval was obtained prior to enrollment of patients in the AllofUs Research Program.

